# Estimation of COVID-19 cases in Mexico accounting for SARS-CoV-2 RT-PCR false negative results

**DOI:** 10.1101/2020.09.17.20197038

**Authors:** Isaac J. Núñez, Pablo F. Belaunzarán-Zamudio, Yanink Caro-Vega

## Abstract

Underestimation of the number of cases during the COVID-19 pandemic has been a constant concern worldwide. Case confirmation is based on identification of SARS-CoV-2 RNA using real time polymerase chain reaction (RT-PCR) in clinical samples. However, these tests have suboptimal sensitivity, especially during the early and late course of infection. Using open data, we estimated that among 1 343 730 people tested in Mexico since February 27^th^, there were 838 377 (95% CL 734 605 – 1 057 164) cases, compared with 604 376 considering only positive tests. ICU admissions and deaths were around 16% and 9% higher than reported. Thus, we show that accounting for the sensitivity of SARS-Cov-2 RT-PCR diagnostic tests is a simple way to improve estimations for the true number of COVID-19 cases in tested people, particularly in high-prevalence populations. This could aid to better inform public health measures and reopening policies.

## Introduction

Around 25 485 000 confirmed cases of COVID-19 have been reported worldwide by country governments by August 31^st^, 2020.^1^ The Mexican government reported 599 560 confirmed cases to that date.^2^ Case confirmation is based on identification of SARS-Cov-2 RNA using real time polymerase chain reaction (RT-PCR) in clinical samples collected through nasopharyngeal or oropharyngeal swabs, saliva, or bronchoscopy. Dealing with the disease has proven extremely challenging for governments and health systems worldwide, partially due to the difficulties in case identification. Various measures have been proposed to reduce the viruses’ impact on the population, most of them rely on case identification for isolation and contact tracing.^3-5^

The World Health Organization (WHO) has highlighted the importance of generalized testing with the goal of early detection, quarantine, and contact tracing. Countries like South Korea and Iceland have been successful in implementing wide-spread testing, case-isolation and contact tracing, keeping the virus under control.^6,7^ In countries where tests are less available, focusing this resource to high risk people was deemed reasonable as a provisional strategy, with the urge to increase testing capacity. Mexico chose a different strategy, and a decision to use testing only for surveillance purposes was made early during the pandemic. Criteria for testing are applied as for the sentinel surveillance system for influenza, and the information provided allegedly used to estimate the total number of infections based on mathematical modelling. As anywhere else, underestimation of the number of cases has been a constant concern.^4-7^

Diagnostic tests rarely, if ever, are completely reliable, and RT-PCR for SARS-CoV-2 is no exception. Specificity almost always nears 100% in this kind of tests, but poor sensitivity has been an issue. Kucirka and collaborators estimated that test sensitivity is highest at the fourth day of symptoms onset (81% [95% confidence limits; 95%CL 71-88%]).^8^ Sensitivity is the lowest during the asymptomatic and late symptomatic periods (eg. 37%, 95%CL 26-49%) at day 21 after symptom onset. Other factors that could influence the accuracy of the test is the type of clinical specimen, severity of infection, and gene targets. A combination of these, and other factors, may account for the underestimation of the number of cases and attributable deaths worldwide.^8-11^ Until screening and diagnostic tests performance are optimized, applying mathematical modelling strategies can aid in estimating more accurately diseases occurrence for surveillance purposes. In this study, we aimed to provide corrected estimates of the number of cases among people that were tested for SARS-Cov-2 in Mexico between February 27^th^ and August 31^th^, 2020 by taking into account the probability of RT-PCR false negative tests results.

## METHODS

### Study setting

In Mexico, the first COVID-19 confirmed case was tested on February 27^th^ and reported on February 28^th^. Community transmission was declared on March 24^th^ and mitigation country-wide measures were announced the same day. Social distancing was urged, and non-essential businesses and activities were suspended, initially until April 14^th^. The testing strategy was also published that day. A case definition was developed, and testing was recommended for one in ten patients seeking care due to a mild case of an influenza like-illness in a limited number of health services previously established to monitor seasonal Influenza, and all of those requiring hospitalization. Also, heavy emphasis on voluntary quarantine if mild symptoms developed, urging people with co-morbidities and other high-risk conditions, such as older age, to search for health care. No accompanying contact tracing measure was spoken of, placing most of the responsibility at the individual level.^12^

### Data sources and selection

We used the SARS-CoV-2 tests open datasets made public since April 12 by the Mexican government in their official coronavirus web page and updated daily.^13^ The datasets include every test done at public, but not private, laboratories. It contains State and Municipality where the test was collected, sociodemographic information, dates of symptoms onset, date the patient was included in the database, ICU admission and death (if occurred) for everyone, with non-traceable, individual key identifiers. We assumed the date the patient was registered in the database was the date of testing, and we will refer to it as such from now on. We included in the analysis all tested individuals registered in the dataset between February 27^th^ and August 31^st^. Patients with pending result, missing identification code, or more than 21 days with symptoms at the moment of the test were excluded.

We analyzed each information according to the date the tests results are reported in the database, regardless of the day it was collected to follow the format of the daily report by the Ministry of Health (See supplementary). This was not possible for people tested before April 12^th^ and already had a result, so their result was included as the baseline count.

### False negative estimation model

We used the method described by Kucirka and collaborators in their mathematical modelling study to calculate the false-negative rate of RT-PCR diagnostic tests. They calculated the sensitivity from day one of infection (assuming symptoms started at the 5^th^ day of infection) until day 21 of infection. Since their estimates end at day 16 of symptoms, we replicated their analysis and estimated sensitivity up to day 21 of symptoms with 95% uncertainty bounds. Sensitivity varied depending on the day after infection, being higher during the symptomatic phase and reaching a maximum of 81% at the fourth day of symptoms (Supp Append Table S1). We used the mean estimate for the graphical representation but repeated the estimation with the upper and lower confidence bounds. Specificity for every test used by the Mexican government is reportedly 100%.^8, 14^

We used the following contingency table as the basis for our analysis:

| Test result | Person has COVID-19 |  | Total |
| --- | --- | --- | --- |
|  | Yes | No |  |
| Positive | a | b | a + b |
| Negative | c | d | c + d |
| Total | a + c | b + d | a + b + c + d |

Given a 100% of specificity, there are no false positive results then b=0, and all the positive results are true positives. From data set, each day we knew the number of true positives ‘a’ and the number of negative tests ‘c+d’. Our interest is to estimate the daily number of false negatives ‘c’. The probability of being false negative ‘p’ is defined in the equation (1):

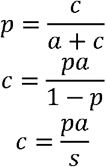

By reproducing Kucirca’s analysis, we know ‘p’ and ‘s’ (1-p) the test sensitivity for each time since symptom initiation reported. At each calendar day, we split individuals in groups, each corresponding to the number of days with symptoms when tested. For example, a day in which 100 people were tested, with 30 presenting on their 6^th^ day and 70 in their 7^th^ day with symptoms, two groups were created, each one with a test sensitivity (*s*) and the number of reported positive tests (*a*). Hence, we applied the equation to each group of every calendar day from February 27^th^ to August 31^st^ and added the false negatives calculated on every group. As the number of true COVID-19 cases is limited by the number of people tested, in case the estimation yielded a higher number of cases then the totality of people tested was used instead.

We also estimated the corrected number of ICU admission and the corrected number of deaths due to COVID-19 by calendar day. Assuming no difference in test precision among disease severity spectrum, we added the product of the proportion of negatives estimated to be false negatives and the number of ICU admissions or deaths among COVID negative patients. We applied the following equation to correct death and ICU admissions:

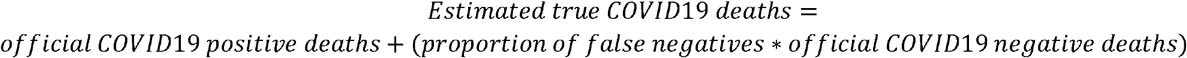

Thus, if in a given day there were 100 deaths and 50 ICU admissions among COVID-19 negative patients, and the estimated false negative proportion using the mean estimate of the test sensitivity was of 0.40, we would add 40 deaths and 20 ICU admissions to the COVID-19 positive group for that particular date.

### Statistical Analysis

Applying results derived from the false negative estimation model on the government official dataset, we estimated the daily corrected number of cases. We performed the analysis at a National level and for each one of the 31 States and the country capital (Ciudad de México, formerly known as Distrito Federal).

To determine if positivity rates could be due to low testing per capita, we calculated Spearman’s rho of positivity rates and the number of tests done per 10 000 habitants by state. State population was obtained from the national statistics and geography institute’s (INEGI) most recent published data.^15^ We also calculated the 7-day moving average of time from symptom onset to testing for the entire study period to determine if this could explain higher false negative rates during certain time periods.

All data analysis was done with R software version 4.0.0. The ethics committee of the Instituto Nacional de Ciencias Médicas y Nutrición Salvador Zubirán reviewed and approved the study. There was no sponsor involved in any step of the study.

## RESULTS

There were 1 343 730 people tested between February 27^th^ and August 31^st^ according to the latest official database (August 31^st^). A detailed explanation of data selection is provided in supplementary materials. We included 1 280 910 patients that had an available result, were tested at less than or 21 days with symptoms and had no missing ID code. Of them, 604 376 (47.2%) were SARS-CoV-2 positive and 676 534 (52.8%) negative (Table 1).

**TABLE 1.** Official and estimated nation-wide COVID-19 cases by date of reporting.

| Variable | Official count | Expected scenario | Best case scenario | Worst case scenario |
| --- | --- | --- | --- | --- |
|  | 1 280 910 | 1 280 910 | 1 280 910 | 1 280 910 |
| Positive tests | 604 376 (47.2%) | 838 377 (65.5%) | 734 605 (57.4%) | 1 057 164 (82.5%) |
| Negative tests | 676 534 (52.8%) | 442 533 (34.5%) | 546 305 (42.6%) | 223 746 (17.5%) |
| Estimated false negative tests | - | 234 001 (18.3%) | 130 229 (10.2%) | 452 788 (35.3%) |
| Overall ICU count | 13 038 | 15 085 | 14 167 | 17 008 |
| Overall Death count | 64 360 | 69 835 | 67 406 | 74 978 |

**Note:** Expected, best and worst-case scenario refer to estimations done with the mean test sensitivity and its upper and lower 95% confidence limits, respectively. Positive tests in the expected, best case and worst-case scenarios include estimated false negatives, while negative tests exclude them.

### Estimated false negatives and corrected COVID-19 cases

We estimated a total of 838 377 (95% CL 734 605 – 1 057 164) positive cases (39% higher than official reported cases) (Figure 1). In our corrected estimates, 50 000 cases were reached by May 11^th^ (May 7^th^-May 14^th^), while the official count reached that number at May 18^th^. Accumulated test positivity rate increased during the study period, being of 14.4% at March 24^th^, 36% at April 12^th^, and 50.1% by July 14^th^. Positivity rate also varied according to state and date (Table 2, Supp Material Figures SE). The states with the most cases were Ciudad de México, Estado de México, Guanajuato, Nuevo León, and Puebla, while those with the highest positivity rate were Veracruz, Oaxaca, Baja California, Quintana Roo and Hidalgo. Spearman’s Rho for test positivity rate (taking false negatives into account) and tests performed per 10 000 people was of −0.41 (−l0.40, −0.41). Time from symptom onset to testing increased over time (Supp Material Figure S1).

**TABLE 2.** Official and estimated confirmed state-wide COVID-19 cases.

| State | Reported cases<br>(positivity rate) | Estimated cases | Estimated<br>positivity rate<br>(95% CL) | Reported<br>ICU<br>admissions | Estimated ICU<br>admissions<br>(95% CL) | Reported<br>deaths | Estimated deaths<br>(95% CL) | Total tests | Tests per<br>10 000 hab |
| --- | --- | --- | --- | --- | --- | --- | --- | --- | --- |
| VERACRUZ | 28 053 (67.3%) | 37 560 (32 986-41 679) | 90.1% (79.1-100%) | 616 | 745 (668-862) | 915 | 1669 (1293-2396) | 41 679 | 51 |
| OAXACA | 13 793 (68.9%) | 17 914 (15 749-20 017) | 89.5% (78.7-100%) | 269 | 339 (297-409) | 434 | 821 (619-1234) | 20 017 | 50 |
| BAJA CALIFORNIA | 17 197 (63.3%) | 23 009 (19 719-27 157) | 84.7% (72.6-100%) | 94 | 139 (102-182) | 1036 | 1804 (1373-2707) | 27 157 | 82 |
| QUINTANA ROO | 10 236 (61.5%) | 13292 (11708-16328) | 79.9% (70.4-98.2%) | 223 | 255 (230-305) | 256 | 397 (315-560) | 16 634 | 111 |
| HIDALGO | 10 095 (60.6%) | 13 208 (11 517-16 365) | 79.3% (69.1-98.2%) | 116 | 135 (118-163) | 348 | 537 (426-752) | 16 666 | 58 |
| SINALOA | 16 191 (57.7%) | 22 092 (19 179-27 795) | 78.7% (68.3-99%) | 597 | 704 (643-804) | 930 | 1626 (1271-2293) | 28 076 | 95 |
| SONORA | 21 210 (58.2%) | 28 618 (24 925-35 917) | 78.5% (68.4-98.5%) | 145 | 180 (161-221) | 884 | 1597 (1234-2314) | 36 460 | 128 |
| CHIAPAS | 6148 (61%) | 7819 (6803-9887) | 77.6% (67.5-98.2%) | 198 | 229 (208-281) | 277 | 526 (361-847) | 10 073 | 19 |
| GUERRERO | 14 659 (59.7%) | 19 038 (16 860-23 298) | 77.5% (68.6-94.8%) | 375 | 436 (394-500) | 604 | 1030 (814-1449) | 24 570 | 70 |
| TABASCO | 28 833 (52.1%) | 41 329 (35 799-52 230) | 74.7% (64.7-94.4%) | 238 | 271 (252-310) | 568 | 878 (730-1155) | 55 312 | 231 |
| QUERETARO | 6623 (57.2%) | 8524 (7362-10 865) | 73.7% (63.6-93.9%) | 91 | 112 (94-153) | 67 | 92 (70-133) | 11 571 | 57 |
| NAYARIT | 5003 (57.9%) | 6260 (5564-7621) | 72.4% (64.4-88.2%) | 163 | 171 (163-190) | 118 | 163 (132-215) | 8645 | 73 |
| COLIMA | 3798 (58.3%) | 4696 (4222-5728) | 72.1% (64.8-88%) | 82 | 92 (85-112) | 106 | 147 (122-206) | 6512 | 92 |
| YUCATÁN | 14 973 (54.1%) | 19 776 (17 272-24 996) | 71.5% (62.4-90.3%) | 159 | 180 (162-208) | 206 | 291 (246-396) | 27 668 | 132 |
| CAMPECHE | 5743 (51.5%) | 7850 (6704-10 048) | 70.4% (60.1-90.1%) | 224 | 249 (231-280) | 112 | 218 (160-329) | 11 152 | 124 |
| PUEBLA | 28 183 (49.8%) | 39 848 (34 192-51 630) | 70.4% (60.4-91.2%) | 448 | 506 (473-583) | 840 | 1277 (1062-1714) | 56 600 | 92 |
| CHIHUAHUA | 7861 (50.4%) | 10 456 (8638-14 257) | 67.1% (55.4-91.5%) | 603 | 744 (638-956) | 456 | 637 (500-909) | 15 586 | 44 |
| ESTADO DE MÉXICO | 49 647 (47.2%) | 67 627 (58 513-87 646) | 64.3% (55.6-83.3%) | 687 | 809 (734-942) | 2924 | 4348 (3595-6065) | 105 239 | 65 |
| SAN LUIS POTOSÍ | 18 343 (47.4%) | 24 616 (21 532-30 699) | 63.7% (55.7-79.4%) | 151 | 180 (159-223) | 336 | 472 (403-596) | 38 664 | 142 |
| TAMAULIPAS | 24 257 (44.2%) | 34 096 (28 952-45 679) | 62.1% (52.7-83.2%) | 129 | 174 (149-247) | 510 | 806 (649-1132) | 54 902 | 160 |
| MORELOS | 5301 (50%) | 6571 (5743-8308) | 61.9% (54.1-78.3%) | 41 | 43 (41-51) | 421 | 920 (523-1714) | 10 611 | 56 |
| NUEVO LEÓN | 29 830 (44.6%) | 40 632 (35 328-52 169) | 60.8% (52.8-78%) | 671 | 796 (731-931) | 160 | 240 (196-329) | 66 869 | 131 |
| COAHUILA | 22 147 (44.2%) | 30 176 (25 862-39 774) | 60.3% (51.6-79.4%) | 58 | 83 (69-115) | 465 | 684 (570-950) | 50 076 | 169 |
| ZACATECAS | 5282 (45.3%) | 6778 (5976-8289) | 58.1% (51.2-71.1%) | 183 | 194 (184-223) | 75 | 87 (77-118) | 11 665 | 74 |
| BAJA CALIFORNIA SUR | 7639 (45.5%) | 9654 (8583-11 933) | 57.4% (51.1-71%) | 70 | 77 (70-90) | 27 | 28 (27-41) | 16 807 | 236 |
| CIUDAD DE MÉXICO | 118 009 (39.6%) | 168 089 (144 621-220 218) | 56.5% (48.6-74%) | 1438 | 1755 (1604-2077) | 2116 | 2829 (2490-3569) | 297 668 | 334 |
| GUANAJUATO | 31 736 (43.1%) | 41 197 (36 692-50 156) | 56% (49.9-68.2%) | 261 | 289 (267-332) | 465 | 600 (532-739) | 73 588 | 126 |
| MICHOACÁN | 15 346 (40.6%) | 21 136 (18 357-26 676) | 55.9% (48.6-70.6%) | 88 | 108 (93-143) | 305 | 394 (351-492) | 37 810 | 82 |
| JALISCO | 20 525 (40.4%) | 27 838 (24 089-35 906) | 54.8% (47.4-70.7%) | 488 | 567 (526-662) | 523 | 698 (612-895) | 50 792 | 65 |
| TLAXCALA | 5571 (34.8%) | 7434 (6396-9485) | 46.5% (40-59.3%) | 441 | 512 (466-599) | 118 | 140 (123-180) | 15 998 | 126 |
| DURANGO | 6414 (34.4%) | 8566 (7378-11209) | 45.9% (39.5-60.1%) | 80 | 86 (80-94) | 35 | 38 (35-44) | 18 663 | 106 |
| AGUASCALIENTES | 5730 (33.4%) | 7699 (6516-10028) | 44.8% (37.9-58.4%) | 28 | 35 (28-56) | 48 | 63 (51-85) | 17 180 | 131 |

**Note: State-wise per-occurrence date analysis, study period February 27<sup>th</sup> – August 31<sup>th</sup>. Ordered by estimated** **positivity rate (highest to lowest)**

**Figure 1.**
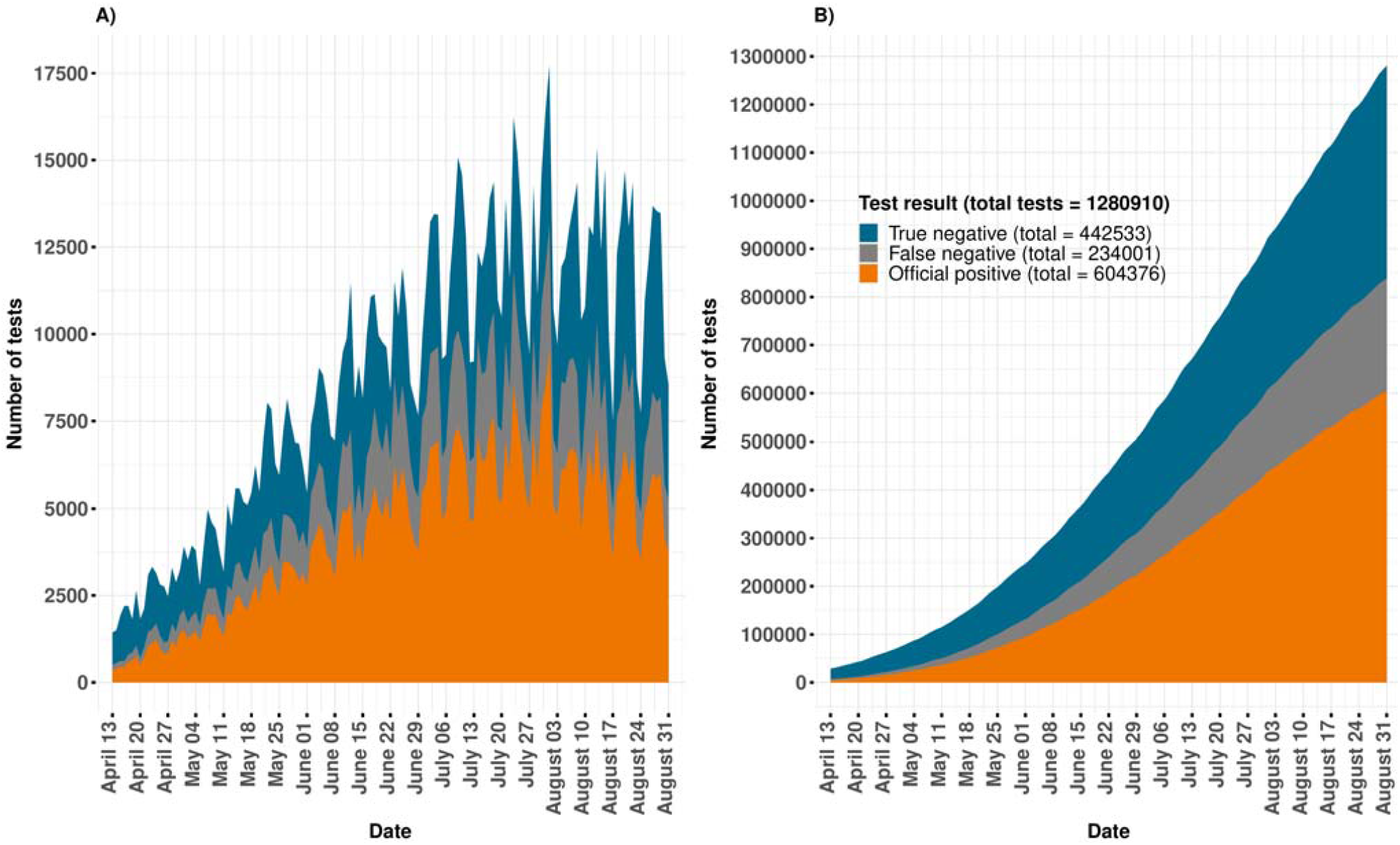
Estimated proportion of tested individuals with a false negative result during the study period.

### Corrected ICU hospitalization and deaths estimates

There were 13 038 ICU admissions and 64 360 deaths among COVID-19 positive patients during the study period (Table 1). The corrected estimate is of 15 085 ICU admissions (14 167 – 17 008) and 69 835 deaths (67 406 – 74 978). (Table 1). The magnitude of difference between official reports and corrected estimates varied between states (Table 2).

## DISCUSSION

In this analysis, we estimated the corrected number of cases of COVID-19, ICU admissions and deaths due to COVID-19 in Mexico accounting for false negatives tests results using test sensitivities previously estimated by Kucirca and colleagues. These were estimated based on the day after the onset of symptoms when patients are tested. We identified that the number of cases of COVID-19 in Mexico based on RT-PCR testing might be almost 40% higher than currently registered, with 95% confidence limits of 21% and 75% depending on the test sensitivity. These differences vary widely by state and period during the pandemic. Accordingly, the corrected number of ICU admission and deaths increased around 16% and 9%, but this increase might be as high as 25% in ICU and 17% in deaths. The magnitude of these differences may require important modifications in preparedness for response, which highlights the importance of accounting for the probability of false negative tests in public health estimations.

### A) represents new daily test results, B) represents accumulated test results

The proportion of false negatives was similar to that found in other studies.^16, 17^ There were high heterogeneity in positivity tests proportions, corrected estimates, and confirmed and corrected estimations of ICU hospitalizations and deaths across States. A modelling study conducted in the United States showed that disease burden varied heavily among counties, both in an optimistic and in a pessimistic scenario.^18^ A recent seroepidemiological study conducted in Spain found considerable heterogeneity in seroprevalence among provinces, with >10% in the most heavily affected ones and less than one percent in the least.^19^ This data is consistent not only with the occurrence of “local epidemics” rather than a nation-wide epidemic; but also the fragmented response in Mexico with some regions faring better than others. On the last point, it may only reflect sound State-centered approaches, with resources being modified according to each states’ needs, though.

We also observed a weak negative correlation between test positivity and tests-per-capita. Considering that testing per-capita is very low in the whole country, this is not surprising. States that have particularly low testing rates are Chiapas, Chihuahua, Oaxaca, Veracruz, and Queretaro. This could be responsible for the high estimated positivity rate. Sonora and Baja California have more tests done per 10 000 habitants, but their estimated positivity rate is still of ∼90%. Most likely the number of cases far outnumber the number of tests, with the relatively small differences in tests-per-capita being apparently inconsequential for the positivity rate. Interestingly, the time from symptom onset to testing did not appear to change considerably over time. This suggests that increasing positivity rates most likely do not derive from changes in time of testing after symptoms onset along time but from high disease burden and insufficient testing worsening over time. Considering that only a small proportion of symptomatic cases who search for healthcare are tested (not even considering asymptomatic individuals who have the virus) the true underestimation of COVID-19 cases can be huge. Given this, the daily number of cases will most likely grow on par with the number of tests.

This means that with current testing capacity it is not possible to grasp the behavior of the pandemic, as the number of tests is so small and the positivity rate so high that it would be fully dependent on them. The World Health Organization recommends a positivity rate lower than 5%, among other criteria, to commence reopening, even if a sentinel system is being used. Mexico is currently far from a safe reopening, and further still if we consider false negatives.^20, 21^

False negatives are accounted for in clinical medicine when a clinician suspects it in a patient that has a negative test but other disease indicators, such as suggestive lung images, that generate a convincing clinical scenario, and acts accordingly.^22^ As we show here, false negatives should also be accounted for in public health estimations, and it is also possible to act accordingly. Places with low prevalence, as in the states with low positivity rate (none in our case) or with massive testing strategies, will have a small amount of cases added to their official counts. This contrasts with the picture of Mexico as a whole, were the worrisome positivity rate increases even more when false negatives are considered.^23, 24^ Currently, a reopening strategy based on a four-coloured traffic light (red, orange, yellow, and green) is being implemented, which assigns each state a colour based on several variables. Test positivity rate is among the criteria for changing the colour. Nonetheless, by not considering false negatives the positivity rate is being underestimated and it is likely this could influence the premature modification of the colour, and thus premature reopening, which could cause a new surge in cases.^25-27^ We observed a one-week delay in reaching a similar number of cases when mean false-negative test is not considered.

This approach as applied for Mexico has several limitations. As only a few symptomatic people are tested we cannot estimate the corrected number of cases among the total population. This would require a vastly higher number of tests. Thus, there should be caution when interpreting our results. Our estimation presents a limited view of how much more cases there are, and thus do not represent the actual number of cases in the country; in any case our corrected estimates are in the conservative side.^14, 28, 29^ We assumed that the date on which the information was captured in the system is the same on which the sample was procured, but there might be a small-time lag between one and another. We also identified limitations related to the diagnostic tests. For instance, there is a wide catalog of SARS-Cov-2 RT-PCR tests currently used in our country. Even if all of them have to be approved by the Institute of Diagnosis and Epidemiological Reference (INDRE) for surveillance purposes and all must comply with the Berlin protocol, we do not know how they compare to each other, and variation in diagnostic accuracy probably exists. We also do not have information on the anatomical test site, be it nasal or oropharyngeal swab, saliva or bronchoscopy sample, which could affect test sensitivity. We do not account for severity of the clinical picture on the test sensitivity. Sensitivity of the test in patients who develop pneumonia and /or critical illness could be higher than in those with less severe disease, and patients with severe disease is overrepresented in the testing strategy followed in Mexico. Even when considering all these limitations, the application of the test performance to correct the number of cases could certainly improve surveillance. The method we used can be easily adapted to other countries or areas. Our analysis can be updated if COVID-19 open data continues to be published, and thus be used to better inform decision making at the National and State level.

## Conclusion

While it may very well be impossible to determine which patients had the false negative tests, taking the test precision into account is an effective way to improve estimations for the number of COVID-19 cases among a tested population, especially when the testing is done mainly in high-prevalence populations. We expect this could aid to better inform public health measures and reopening policies.

## Data Availability

All code utilized will be available at https://github.com/isaac-nunez/covid_19_fn_estimates_mexico and all datasets are available at the official government COVID-19 webpage.

## Acknowledgements

We would like to thank all the health-workers in our country for their tremendous work in caring for patients during this pandemic. Also, we thank the people behind the surveillance of the pandemic, as without them this work could not have come to be.

## Declaration of interests

All authors state they have no conflict of interests.

## Author contributions

Isaac J. Núñez: idea conception, literature search, study design, data curation, data analysis, data interpretation, writing Pablo F. Belaunzarán-Zamudio: literature search, study design, data analysis and interpretation, writing Yanink Caro-Vega: literature search, study design, data curation, data analysis, data interpretation, writing

## Data sharing

All code utilized is available at https://github.com/isaac-nunez/covid_19_fn_estimates_mexico and all datasets are available at the official government COVID-19 webpage.^13^

## Notes

### Competing Interest Statement

The authors have declared no competing interest.

### Funding Statement

We have received no funding for this study.

### Author Declarations

This study was approved by the ethics committee of the Instituto Nacional de Ciencias Medicas y Nutricion Salvador Zubiran.

